# Using deep learning methods to shorten acquisition time in children’s renal cortical imaging

**DOI:** 10.1101/2025.08.11.25333418

**Authors:** Cui Gan, Pei Niu, Boyang Pan, Xiaoxiao Chen, Libo Xu, Kanlei Huang, Hailin Chen, Qunli Wang, Lin Ding, Yafu Yin, Shuqi Wu, Nan-Jie Gong

## Abstract

**Purpose:** This study evaluates the capability of diffusion-based generative models to reconstruct diagnostic-quality renal cortical images from reduced-acquisition-time pediatric 99mTc-DMSA scintigraphy.

**Materials and Methods:** A prospective study was conducted on 99mTc-DMSA scintigraphic data from consecutive pediatric patients with suspected urinary tract infections (UTIs) acquired between November 2023 and October 2024. A diffusion model SR3 was trained to reconstruct standard-quality images from simulated reduced-count data. Performance was benchmarked against U-Net, U^2^-Net, Restormer, and a Poisson-based variant of SR3 (PoissonSR3). Quantitative assessment employed peak signal-to-noise ratio (PSNR), structural similarity index (SSIM), Fréchet inception distance (FID), and learned perceptual image patch similarity (LPIPS). Renal contrast and anatomic fidelity were assessed using the target-to-background ratio (TBR) and the Dice similarity coefficient respectively. Wilcoxon signed-rank tests were used for statistical analysis.

**Results:** The training cohort comprised 94 participants (mean age 5.16±3.90 years; 48 male) with corresponding Poisson-downsampled images, while the test cohort included 36 patients (mean age 6.39±3.16 years; 14 male). SR3 outperformed all models, achieving the highest PSNR (30.976±2.863, P<.001), SSIM (0.760±0.064, P<.001), FID (25.687±16.223, P<.001), and LPIPS (0.055±0.022, P<.001). Further, SR3 maintained excellent renal contrast (TBR: left kidney 7.333±2.176; right kidney 7.156±1.808) and anatomical consistency (Dice coefficient: left kidney 0.749±0.200; right kidney 0.745±0.176), representing significant improvements over the fast scan (all P < .001). While Restormer, U-Net, and PoissonSR3 showed statistically significant improvements across all metrics, U^2^-Net exhibited limited improvement restricted to SSIM and left kidney TBR (P < .001).

**Conclusion:** SR3 enables high-quality reconstruction of 99mTc-DMSA images from 4-fold accelerated acquisitions, demonstrating potential for substantial reduction in imaging duration while preserving both diagnostic image quality and renal anatomical integrity.

## Introduction

Urinary tract infection (UTI) [1] is a common condition in pediatric patients, affecting around 1.7% of males and 8.4% of females before the age of 7 years. Clinical symptoms of UTI encompass flank pain with fever, urgency and frequent voiding, incontinence, and dysuria among older children, with other long-term renal damage [2, 3]. Dimercaptosuccinic acid (DMSA) scintigraphy is widely advocated for evaluating renal cortical defects, detecting parenchymal damage, and assessing the presence of structural abnormalities, making precise DMSA scintigraphy crucial for improving patient outcomes and guiding clinical interventions [4].

However, clinical challenges persist in pediatric DMSA scintigraphy for UTI diagnosis[5]. Firstly, The procedure is prolonged (typically >20 minutes) and requires absolute stillness, which is difficult for children to maintain, increasing motion artifact risk[6]. To improve patient compliance and ensure image quality, mild sedation is frequently administered to infants and uncooperative young children [7]. This carries risks, including hypoventilation, apnea, airway obstruction, and even cardiopulmonary arrest, along with associated morbidity and mortality[8].

Secondly, to reduce radiation risks in infants and young children, low-dose protocols utilize minimal radiopharmaceutical activity [5]. However, fewer injected counts result in lower image contrast and poorer diagnostic clarity [9]. There is also a striving to shorten acquisition protocols, which can help limit motion and reduce the need for sedation. Nevertheless, maintaining a sufficient SNR arises as a challenge since faster scans collect fewer signals, which further degrades image quality [10,11]. In this context, pediatric DMSA imaging necessitates reconstruction methods capable of preserving high image quality under reduced-dose and shortened-acquisition-time conditions [12].

Various deep learning (DL) techniques have been proposed to reconstruct high-quality images from low-count or fast acquisitions [13].These techniques exceed Monte Carlo-based iterative image reconstruction algorithms by achieving equal quality with significantly less time[19]. Previous fast SPECT reconstruction algorithms have demonstrated robust performance in cardiac [14–16] and skeletal [17,18] imaging, achieving significantly improved image quality compared to conventional fast-scan results. U-Net models and Generative Adversarial Networks (GANs) are two commonly adopted architecture in these studies. Isaac Shiri et al. applied Ordered Subsets Expectation Maximization (OSEM) algorithm and a residual neural network to reconstructing SPECT images from half-time acquisitions[15]. Sun et al. proposed a Pix2Pix GAN method in denoising low dose MP SPECT images[16]. While these methods improved noise suppression, Convolutional Neural Networks(CNN)- and Residual Net (ResNet)-based models often produce over-smoothed results, and GANs are limited by training instability and parameter sensitivity [19]. Denoising diffusion probabilistic models (DDPM) [20] have shown promise in medical image restoration, which is proven to be capable of generating images with superior boundary detail preservation while achieving comparable or enhanced overall quality relative to prior approaches[21-23].

Super-Resolution via Repeated Refinement (SR3) is a U-Net based diffusion model, featuring a minimized well-defined loss function, less artifacts in regions with fine-grained texture and a constant number of inference steps regardless of output resolution. It has demonstrated impressive results in natural image super-resolution through iterative stochastic refinement, with a wide range of magnification factors [25]. However, its use in nuclear medicine, particularly in pediatric renal SPECT, remains largely unexplored.

This study evaluates the feasibility of using SR3 to reconstruct pediatric renal cortical DMSA images acquired at one-fourth the standard count level. To better account for the photon statistics of low-count nuclear imaging, the Gaussian noise in the original diffusion process is replaced with a Poisson downsample, resulting in a modified variant termed PoissonSR3. Our study presents the following innovative contributions: 1) To our knowledge, this study represents the first systematic effort to optimize kidney scintigraphy imaging with the application of diffusion models. 2) We proposed two quantitative metrics: Dice Similarity Coefficient (DSC) and Target-to-Background Ratio (TBR) to evaluate renal contrast and anatomical consistency, establishing new evaluation standards for renal scintigraphy.

## Materials and Methods

### 2.1 Patient selection

This prospective study was approved by the Ethics Committee of Xin Hua Hospital Affiliated to Shanghai Jiaotong University School of Medicine (Approval No. XHEC-D-2025-116). Informed consent was obtained from the parents or legal guardians of all pediatric participants prior to DMSA renal scintigraphy. From November 2023 to October 2024, a total of 185 children presenting with UTIs or vesicoureteral reflux as the main complaints underwent DMSA renal scan in our department. Among these cases, 94 were randomly selected for model training, and an additional 36 cases were selected as the test set for further analysis. The demographic and clinical characteristics of the enrolled patients are summarized in Table 1. The selection criteria were as follows: (a) Age ≤ 14 years old; (b) Children with or without of fever ≥38.3°C, elevated C-reactive protein, abnormal urine sediment (pyuria, bacteriuria, hematuria), or positive urine culture (single colony count ≥105 / ml); (c) Agree and be able to cooperate to complete the entire examination.

**Table 1.** Demographic characteristics of patients.

|  | Train Set | Test Set |
| --- | --- | --- |
| No. of participants | 94 | 36 |
| Age at examination | $5.16 \pm 3.90$ | $6.39 \pm 3.16$ |
| Gender |  |  |
| Male | 48 | 14 |
| Female | 46 | 22 |

The exclusion criteria are as follows: (a) Presence of a solitary kidney; (b) Coexisting urinary tract anomalies, such as secondary vesicoureteral reflux, neurogenic bladder, posterior urethral valve, etc; (c) Incomplete or poor-quality imaging due to motion artifacts or failure to complete the scan.

### 2.2 Scanning protocols

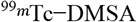 was intravenously injected at a dose of 0.05 mCi/kg, and routine imaging was performed 2-3 hours after injection. Scintigraphy images were obtained using GE discovery nm630 SPECT. The scanning parameters are set as follows: low-energy and high-energy resolution collimators, energy peak 140kev, window width 20%, acquisition matrix 512 × 512, zoom 1.5. Each patient underwent sequential two acquisition: an initial standard scan collecting 600K counts as the reference standard, followed by an accelerated scan acquiring 150K counts.. The children were in a supine position. The detector’s field of view covered the abdominal cavity and pelvic cavity, and the posterior position, anterior position, left posterior oblique position and right posterior oblique position were obtained respectively. All the scans were performed sequentially on the same imaging system to ensure consistency in patient positioning and acquisition parameters.

### 2.3 Diffusion model

SR3 is built upon the Markov chain–based denoising framework of DDPM. In this architecture, low-quality images are provided as conditional inputs at each step of the reverse diffusion process. The model receives both the current noisy image and the corresponding low-count condition, enabling the progressive generation of high-quality reconstructions.

#### 2.4.1 SR3 principle

Input-output image pairs denoted as 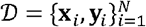, SR3 learns a parametric approximation to *p* (***y***|***x***). SR3 mainly includes forward process *q* and reverse process *p*, where the forward process is the gradual addition of Gaussian noise to high-quality images, and the reverse process is the inverse estimation of the noise vector for each step. Ultimately, SR3 reconstructs the target image ***y***_0_ from a pure noise image **y**_*T*_ through *T* iterative refinement steps.

The Markov forward process *q* of each step is:

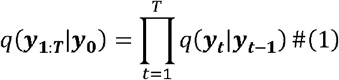

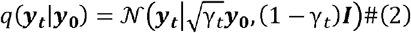

where 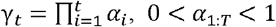 are hyper-parameters, with. In this work, *α*_*t*_*=*1−*β*_*t*_ In this work, *β*_*t*_ values are fixed, following the standard noise schedule design.

Reverse process *p* continuity optimize a neural denoising model 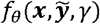 that takes image *x* and noisy target image 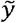 as input source. The output of 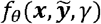 is the noise estimation of each step. Then each iterative refinement under reverse process *p*:

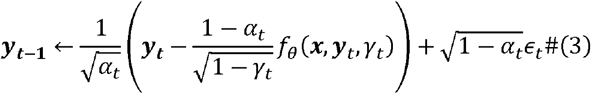

where *∈*_*t* ∼_𝒩 (**0**,***I***). Reverse process *p* is also defined as reverse Markovian process.

#### 2.4.2 SR3 Model Architecture

The SR3 adopts a U-Net structure similar to DDPM but incorporates scaled skip connections, additional residual blocks, and increased channel multipliers at different resolutions. The SR3 U-Net architecture is shown in Fig. 1.

**Fig. 1.**
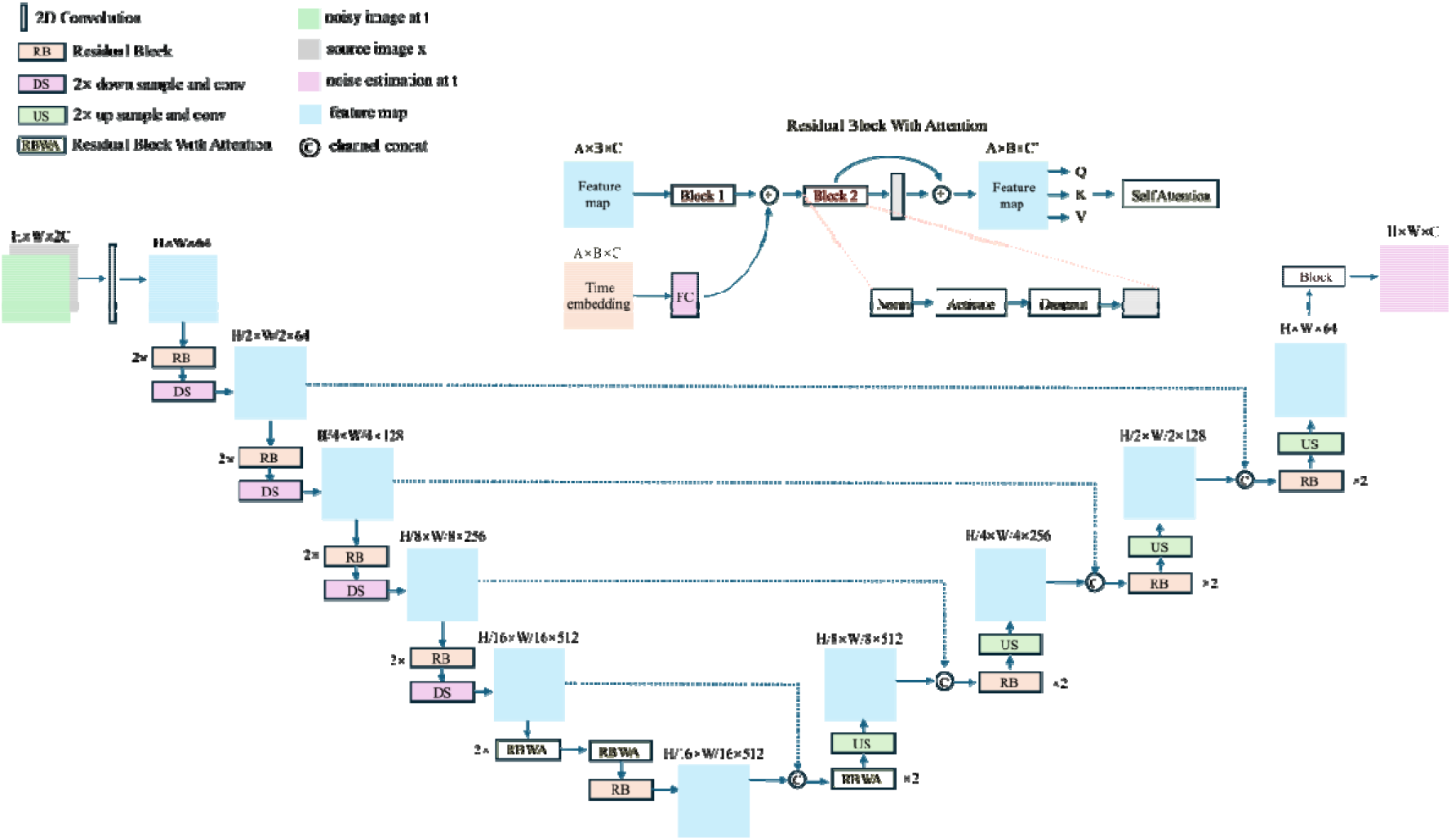
Detailed SR3 model architecture

#### 2.4.3 Optimization

The objective function of the neural network trained in the denoising process p to get noise vector refinement steps, if the noise diffusion process is regarded as an a posteriori estimate of a stationary denoising process, the objective function can be simplified as:

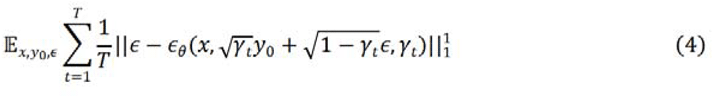

### 2.5 Training details

The training set includes data from 94 participants, and the test set includes 36 participants. For the training set, simulated fast scans were generated from standard scans using Poisson downsample, so as to approximate the photon noise characteristics of low-count acquisitions and enhance data diversity.

The SR3 diffusion model is implemented using a U-Net backbone with 64 initial channels and channel multipliers of [1, 2, 4, 8, 8] across five resolution levels. A self-attention mechanism is applied at the 16×16 feature map level. The model receives a two-channel input composed of a noisy image and the corresponding fast scan image, and produces a single-channel denoised output. The diffusion process consists of 200 timesteps with a linear noise schedule, where β increases from 1×10^−6^ to 1×10^−2^.

Training is performed using the Adam optimizer with β_1_set to 0.9 and β_2_set to 0.999. The learning rate is fixed at 1×10^−4^ for a total of 100,000 iterations. A batch size of 8 is used to balance memory usage and training stability. All models are implemented in PyTorch and trained on an NVIDIA GeForce RTX 3090 GPU. To ensure fair comparison, all models are trained from random initialization with no pretraining or fine-tuning.

### 2.6 Evaluation Metrics

#### 2.6.1 Quantitative metrics

Due to low radiotracer counts and gamma camera efficiency, children’s renal cortical images have inherently low signal-to-noise ratio (SNR), reducing the effectiveness of peak signal-to-noise ratio (PSNR) and structural similarity index (SSIM) [26] for quality assessment. In contrast, Fréchet inception distance (FID) [27] and learned perceptual image patch similarity (LPIPS) [28] better capture perceptual similarity by modelling high-level structural features. Therefore, PSNR, SSIM, FID, and LPIPS are jointly adopted for comprehensive image quality evaluation. The metrics are defined as follows:

PSNR measures the peak pixel-level fidelity by comparing intensity differences between reconstructed and reference images.

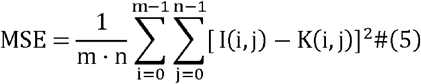

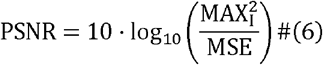

SSIM evaluates structural similarity based on luminance, contrast, and texture consistency between images.

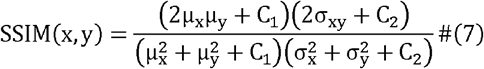

where C_l_ and C_2_ are small constants to stabilize division.

FID quantifies distributional differences in deep feature space using embeddings from a pre-trained Inception-V3 network.

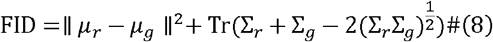

where (*μ*_*r*_, ·_*r*_) and (*μ* _*g*_, ·_*g*_) denote mean and covariance of real and generated features, *T*_*r*_(·) denotes the matrix trace operator. Lower values indicate superior perceptual fidelity.

LPIPS computes perceptual similarity by measuring deep feature distances between image patches using a pre-trained VGG-16 network.

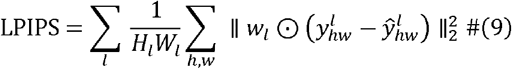

where 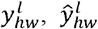 are activations in layer *l*, and *w*_*i*_ are learned weights, *H*_*i*_ and *W* denote the height and width of the *l* layer activation map.

#### 2.6.2 Kidney Contrast and Structural Consistency Metrics

For the standard scans, a threshold-based segmentation method is applied to isolate the left and right kidneys. A count threshold is set at 10% of the maximum counts within the renal region to define the kidney boundaries. The segmented regions are then analyzed to compute the following parameters:

Target-to-Background Ratio. Computed as the ratio of the average counts within each kidney to the average counts in a background region.

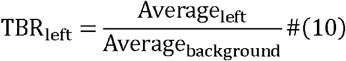

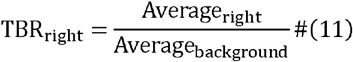

The background region was manually contoured beneath the kidney in each standard scan.

Dice Similarity Coefficient (DSC). Spatial overlap between reconstructed and ground truth segmentations was quantified using DSC, computed separately for each kidney:

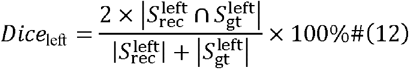

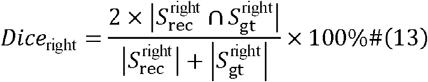

Where *S*_gt_ and *S*_rec_ denote ground truth and reconstructed segmentations respectively.

### 2.7 Statistical Analysis

Statistical analysis is conducted using Python (version 3.11) with the SciPy library (version 1.15.2). Quantitative metrics, including PSNR, SSIM, FID, LPIPS, Dice, and TBR, are compared between reconstruction methods and fast scan images using the Wilcoxon signed-rank test. For all analyses, P < .05 is considered statistically significant difference.

## Results

### 3.1 Objective image analysis

The diffusion model SR3 is compared with three widely used deep learning methods, including U-Net [29], U^2^-Net [30], and Restormer [31]. Additionally, inspired by the conceptual approaches proposed in Cold Diffusion [32] and PDPM [33], the Gaussian diffusion process is modified into a Poisson diffusion process, termed PoissonSR3. Specifically, in the forward process, a stepwise Poisson downsample is applied to standard scan images in order to simulate the degradation consistent with 4× fast scan acquisitions. In the reverse process, standard scan images are gradually reconstructed from the real 4× fast scan inputs by the model.

#### 3.3.1 Quantitative analysis

Table 2 summarizes the quantitative results comparing 4× fast scans and reconstruction outputs. The fast scan yields degraded image quality (PSNR: 25.158±1.778dB; SSIM: 0.506±0.093; FID: 53.234±19.171; LPIPS: 0.133±0.024). Among the methods, SR3 achieves the best overall performance, significantly outperforming the fast scan across all metrics (PSNR: 30.976±2.863dB; SSIM: 0.760±0.064; FID: 25.687±16.223; LPIPS: 0.055±0.022, all P < .001). Restormer and PoissonSR3 significantly improve all four metrics (all P < .001). U-Net also improves across all metrics, with P < .001 in all metrics except for FID (P = .003). In contrast, U^2^-Net fails to improve PSNR, FID, or LPIPS (all P > .99), with significance observed only in SSIM (0.540±0.064, P < .001).

**Table 2.** PSNR, SSIM, FID and LPIPS of the fast scan and outputs of different deep learning methods.

|  | PSNR | SSIM | FID | LPIPS |
| --- | --- | --- | --- | --- |
|  | Mean ±SD | Mean ±SD | Mean ±SD | Mean ±SD |
| Fast Scan(4x) | 25.158±1.778 | 0.506±0.093 | 53.234±19.171 | 0.133±0.024 |
| U-Net | 27.954±1.977 | 0.578±0.091 | 42.228±22.688 | 0.076±0.021 |
| U<sup>2</sup>-Net | 21.435±1.024 | 0.540±0.064 | 274.971±63.549 | 0.261±0.039 |
| Restormer | 29.440±3.507 | 0.671±0.137 | 29.634±16.709 | 0.055±0.023 |
| PoissonSR3 | 30.926±4.012 | 0.736±0.092 | 31.738±32.704 | 0.070±0.058 |
| SR3 | **30.976±2.863** | **0.760±0.064** | **25.687±16.223** | **0.055±0.022** |

Based on the results, U-Net, U^2^-Net, and Restormer exhibit limited consistency in enhancing low-count renal cortical images, while PoissonSR3 presents variable performance across cases. As shown in Fig. 2, U^2^-Net captures the general kidney shape but lacks detail and deviates from the standard scan. U-Net and Restormer produce images closer to the standard, but still show intensity inconsistencies. Diffusion models, PoissonSR3 and SR3, better preserve fine structures via progressive denoising. PoissonSR3 accumulates errors during reverse diffusion, requiring averaging to reduce them, which leads to texture smoothing. In contrast, SR3 benefits from the self-correcting nature of Gaussian noise prediction, preserving contrast and anatomical structure. As illustrated in Fig. 3, SR3 most closely matches the standard scan, maintaining structural and visual fidelity. PoissonSR3 retains the main features but shows patch artifacts from noise accumulation. U^2^-Net performs the worst, with distorted morphology and blurred details. U-Net and Restormer produce plausible but less detailed images than SR3.

**Fig. 2.**
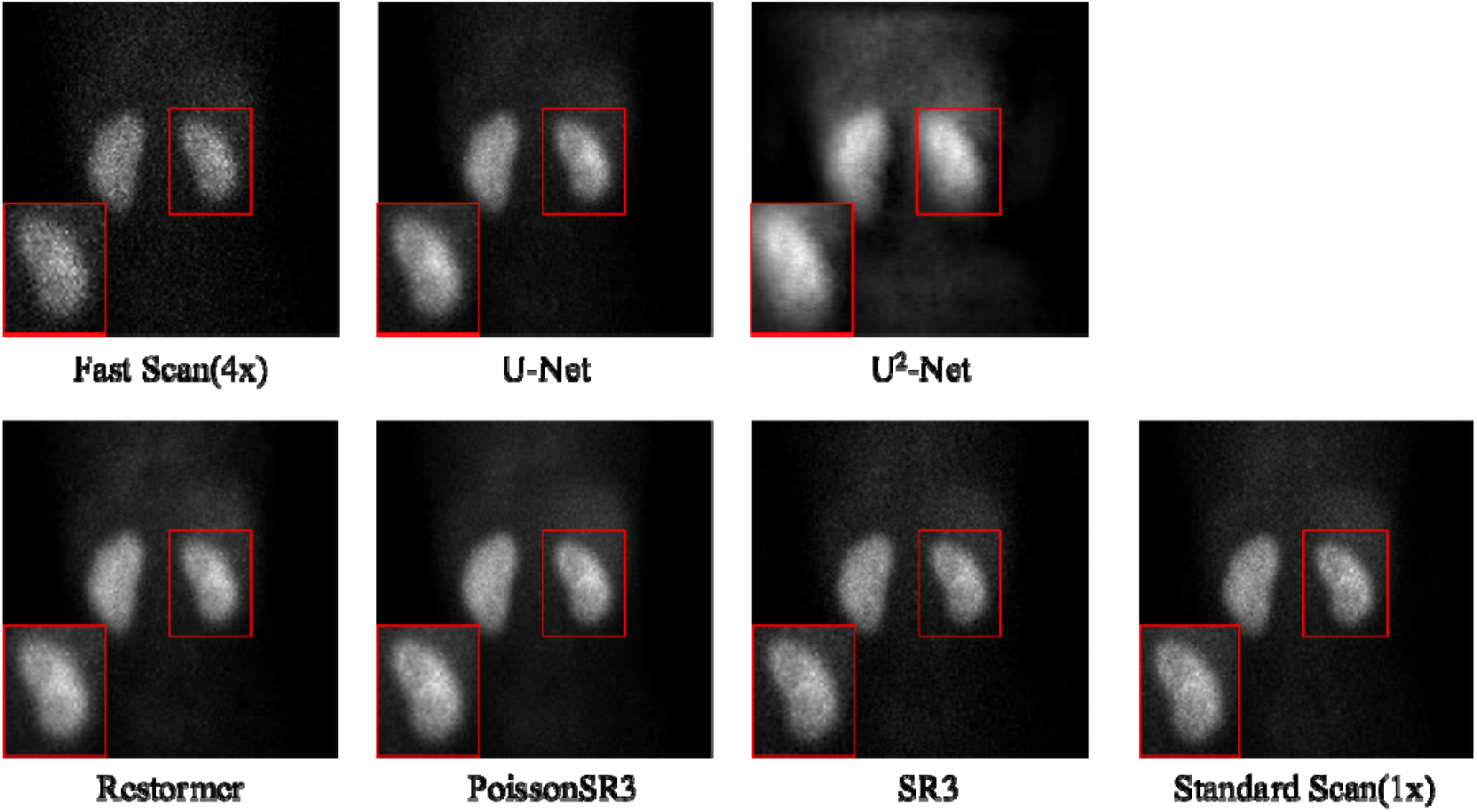
Visualization of children’s right renal cortical imaging. SR3 outperforms all other methods and yields restored high quality with fidelity. In the lower-left corner is the magnified result, which provides enhanced detail visualization.

**Fig. 3.**
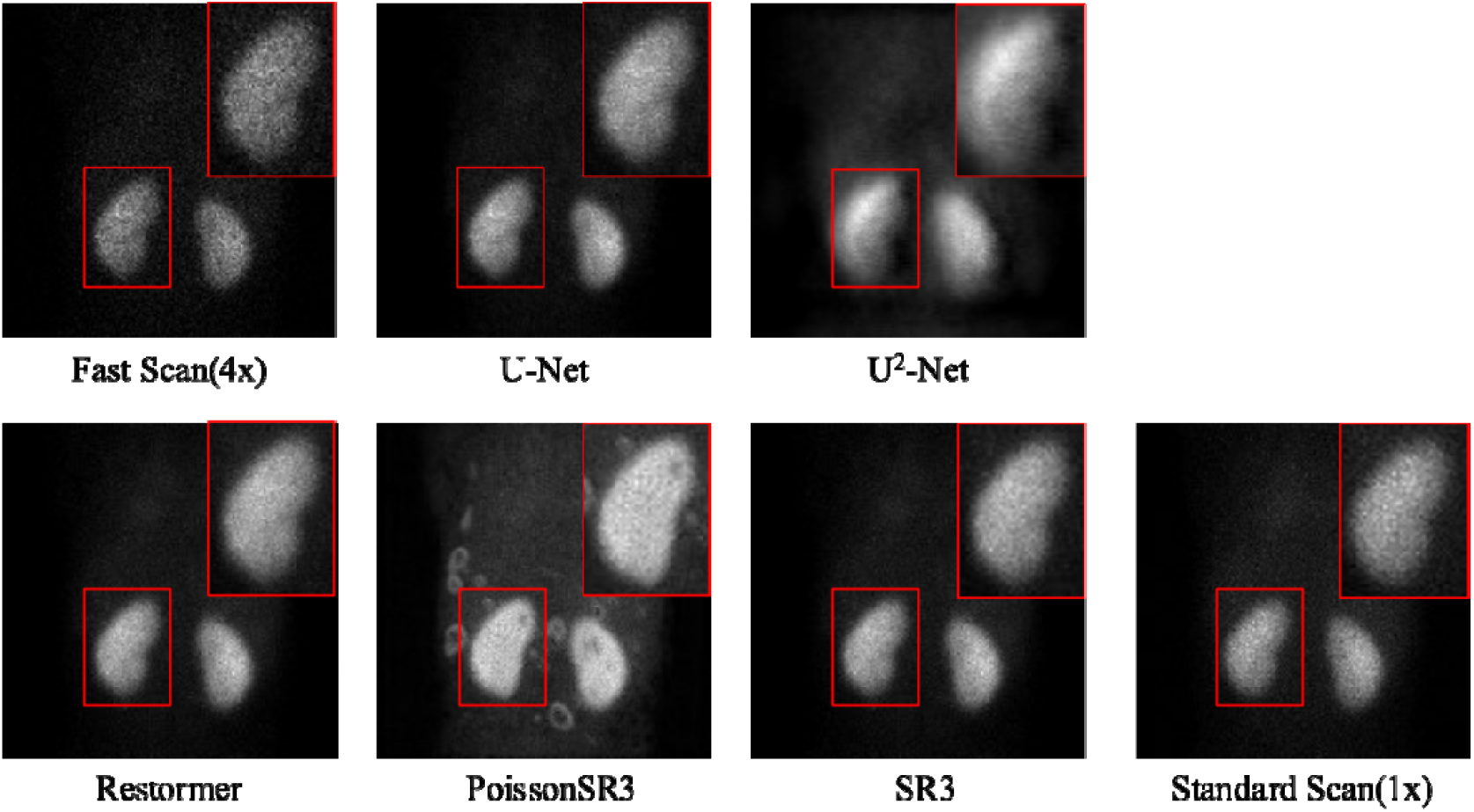
Visualization of children’s left renal cortical imaging. SR3 outperforms all other methods and yields restored high quality with fidelity. In the lower-left corner is the magnified result, which provides enhanced detail visualization.

#### 3.1.2 Kidney Contrast and Structural Consistency Analysis

Table 3 summarizes the contrast and structural consistency performance based on TBR and Dice metrics. The 4× fast scan yields the lowest performance (TBR: 4.206±2.282 left, 4.742±2.178 right; Dice: 0.397±0.223 left, 0.441±0.211 right). SR3 achieves the best results with significant improvements compared with the 4× fast scan(TBR: 7.333±2.176 left, 7.156±1.808 right; Dice: 0.749±0.200 left, 0.745±0.176 right; all P < .001). PoissonSR3, Restormer and U-Net also shows strong improvements (all P < .001), but slightly lower than SR3. U^2^-Net shows the weakest performance, with limited TBR improvement (5.435±1.298 left, P < .001; 4.711±0.883 right, P = .346) and low Dice scores (0.277±0.092 left, P > .99; 0.211±0.078 right, P > .99), failing to achieve significance in most measures.

**Table 3.** TBR and Dice of the fast scan and outputs of different deep learning methods.

|  | TBR<sub>left</sub> | TBR<sub>right</sub> | Dice<sub>left</sub> | Dice<sub>right</sub> |
| --- | --- | --- | --- | --- |
| | Mean ±SD | Mean ±SD | Mean ±SD | Mean ±SD |
| Fast Scan(4x) | 4.206±2.282 | 4.742±2.178 | 0.397±0.223 | 0.441±0.211 |
| U-Net | 6.997±2.371 | 6.800±1.874 | 0.673±0.199 | 0.669±0.182 |
| U<sup>2</sup>-Net | 5.435±1.317 | 4.711±0.896 | 0.277±0.093 | 0.211±0.079 |
| Restormer | 6.934±1.846 | 6.854±1.670 | 0.721±0.205 | 0.736±0.173 |
| PoissonSR3 | 6.996±2.044 | 7.030±1.725 | 0.740±0.199 | 0.723±0.189 |
| SR3 | **7.333±2.176** | **7.156±1.808** | **0.749±0.200** | **0.745±0.176** |

The segmentation visualization results are presented in Fig. 4 and Fig. 5. The outputs of U-Net, U^2^-Net, Restormer, and PoissonSR3 all exhibit varying extent of deviation from the standard scan. In cases with bilaterally consistent renal morphology as illustrated in Fig. 4, U^2^-Net shows noticeable discrepancies in both exterior and interior regions of the kidney. U-Net, Restormer, and PoissonSR3 generally preserve the external kidney boundaries, but inconsistencies remain in the accuracy of delineating internal kidney structures. In contrast, SR3 maintains consistent segmentation for both internal and external regions, indicating its suitability for accurate image reconstruction under reduced acquisition conditions in pediatric renal cortical imaging. In cases involving abnormal renal morphology as illustrated in Fig. 5, U^2^-Net fails to retain structural consistency, whereas U-Net, Restormer, and PoissonSR3 exhibit minor inconsistencies in internal kidney structures compared with the standard scan. SR3 generates segmentation results that most closely align with the standard scan, demonstrating enhanced robustness in challenging renal structural conditions.

**Fig. 4.**
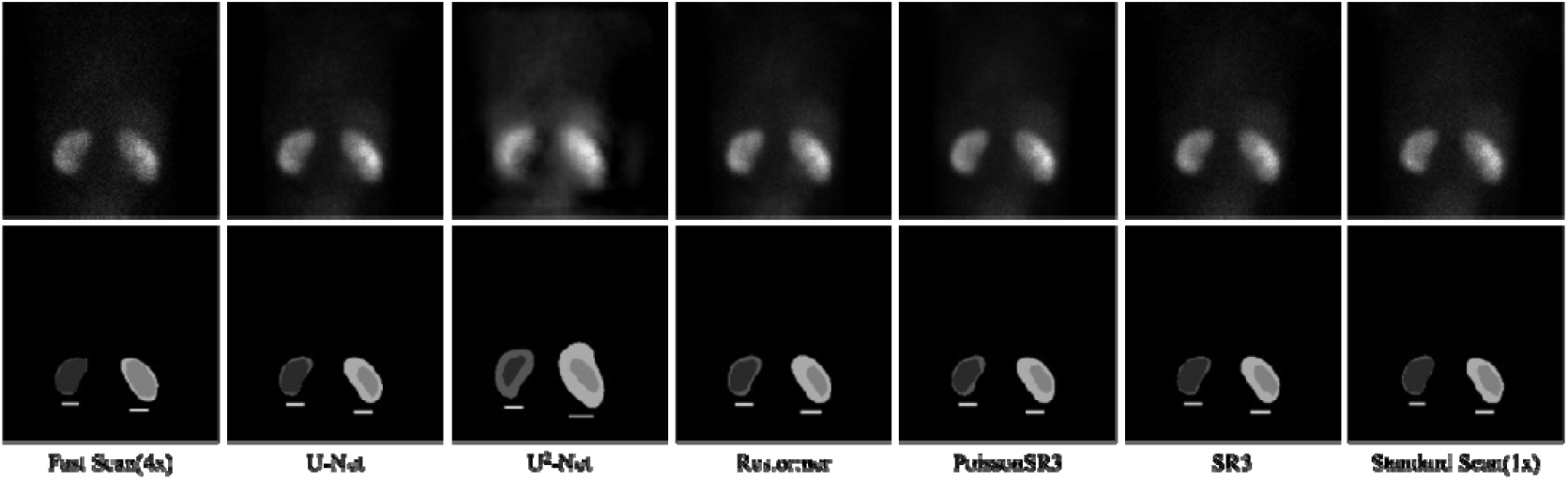
Visualization of segmentation result in cases with bilaterally consistent renal morphology. SR3 outperforms all other methods and yields restored high quality with fidelity. Zoom in for better observe the details.

**Fig. 5.**
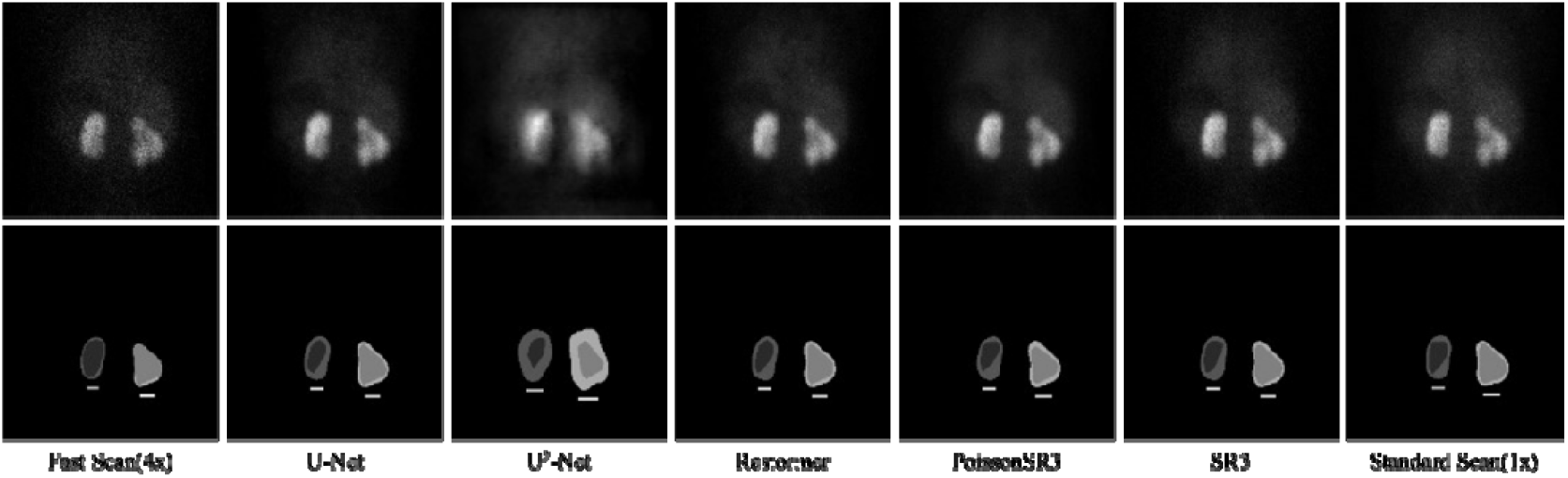
Visualization of segmentation result in cases with abnormal or asymmetric renal morphology. SR3 outperforms all other methods and yields restored high quality with fidelity. Zoom in for better observe the details.

## Discussion

Prolonged acquisition time in pediatric renal cortical Dimercaptosuccinic acid scintigraphy increases the risk of motion artifacts, thus necessitating sedation in infants and uncooperative children. To address this issue, a diffusion model SR3 is trained and evaluated for reconstructing high-quality renal cortical images under accelerated acquisition protocols, aiming to reduce scan time while preserving diagnostic integrity.

Compared with widely used deep learning models, including U-Net, U^2^-Net, and Restormer, SR3 consistently demonstrates superior performance across image fidelity metrics (PSNR, SSIM), perceptual quality metrics (FID, LPIPS) and structural consistency metrics (DSC, TBR). Compared with fast scans, it achieves significant improvements (P< .001) in pixel-wise fidelity and perceptual quality, with reduced distortion and improved structural integrity. SR3 also achieves the best results in maintaining anatomical structural consistency, as demonstrated by improved TBR and Dice (all P< .001). These findings indicate that SR3 not only improves visual realism but also preserves clinically relevant structural details in low-count SPECT images.

In addition to SR3, a modified model referred to as PoissonSR3 is developed in this study to better simulate the photon statistics of low-count SPECT acquisitions. This model replaces the Gaussian noise in the diffusion process with Poisson downsample, aiming to provide physically realistic degradation and restoration pathways. Although PoissonSR3 preserves image contrast reasonably well and shows moderate improvements in perceptual metrics, its performance is less consistent than SR3, particularly in kidney structural consistency and visual detail restoration. This limitation could result from error accumulation during the reverse process, which leads to instability in the generation process. Unlike the continuous Gaussian assumption that can be dealt with smoother optimization and stable denoising trajectories, the non-Gaussian and count-dependent nature of the Poisson process introduces additional variability. As a result, PoissonSR3 often produces patch-like artifacts or blurring, limiting its effectiveness in high-fidelity reconstruction. While PoissonSR3 introduces more realistic noise modeling through its discrete sampling approach, we observe opportunities to further enhance the reverse process trajectory design by optimizing sampling strategies to simultaneously preserve the method’s realistic noise characteristics while maintaining reconstruction stability and fidelity.

In terms of image quality evaluation, we incorporate perceptual metrics (FID, LPIPS) in addition to traditional quantitative measures (PSNR, SSIM), providing a more comprehensive assessment that better aligns with human visual perception. Furthermore, this study introduces a novel methodological approach by implementing DSC and TBR as structural evaluation metrics, specifically tailored to the diagnostic workflow and inherent characteristics of renal scintigraphy. Our model scored significantly higher (p < .001) in all metrics compared with other models, demonstrating its comprehensive ability to enhance the quality of scintigraphy image.

While our quantitative metrics assess image quality and structural consistency, they have inherent limitations for evaluating clinical utility., future work should incorporate subjective rating from experienced nuclear physicians to evaluate the interpretability and practicability of the generated images in clinical settings. Besides, the prospective nature of the data collection from a single centre and the relatively small sample size may have led to biased results. We expect to conduct a multicenter study in the future, adding to the robustness of our model.

In conclusion, this study demonstrates that SR3 provides a robust and effective solution for shortening acquisition time in children’s renal cortical imaging. By enabling high-quality image reconstruction from 4× fast scans, SR3 supports safer and faster imaging protocols and improves pediatric patient comfort and compliance.

## Data Availability

All data produced in the present study are available upon reasonable request to the authors.

## Statements and Declarations

### Author Contributions

All authors contributed to the study conception and design.Data collection was performed by Niu Pei, Kanlei Huang, Hailin Chen, Qunli Wang and Lin Ding. Data analysis was performed by Pei Niu. Clinical Protocol design was performed by Yafu Yin and Shuqi Wu. Technical implementation was conducted by Cui Gan and Boyang Pan. Nan-jie Gong was responsible for Technical and research protocol design. Libo Xu and Xiaoxiao Chen were responsible for manuscript revision. All authors have commented on previous versions of the manuscript. All authors read and approved the final manuscript.

### Ethics approval

All procedures performed in our study were in accordance with the ethical standards of the institutional and/or national research committee and with the 1964 Helsinki declaration and its later amendments or comparable ethical standards. This prospective study was approved by the Ethics Committee of Xin Hua Hospital Affiliated to Shanghai Jiaotong University School of Medicine (Approval No. XHEC-D-2025-116).

### Consent to participate

Informed consent was obtained from all individual participants included in the study.

### Consent to publish

The authors affirm that human research participants provided informed consent for publication of the images in Figure(s) 2, 3, 4 and 5

